# SUMMIT-FA: A new resource for improved transcriptome imputation using functional annotations

**DOI:** 10.1101/2023.02.02.23285208

**Authors:** Hunter J. Melton, Zichen Zhang, Chong Wu

## Abstract

Transcriptome-wide association studies (TWAS) integrate gene expression prediction models and genome-wide association studies (GWAS) to identify gene–trait associations. The power of TWAS is determined by the sample size of GWAS and the accuracy of the expression prediction model. Here, we present a new method, the Summary-level Unified Method for Modeling Integrated Transcriptome using Functional Annotations (SUMMIT-FA), that improves the accuracy of gene expression prediction by leveraging functional annotation resources and a large expression quantitative trait loci (eQTL) summary-level dataset. We build gene expression prediction models using SUMMIT-FA with a comprehensive functional database MACIE and the eQTL summary-level data from the eQTLGen consortium. By applying the resulting models to GWASs for 24 complex traits and exploring it through a simulation study, we show that SUMMIT-FA improves the accuracy of gene expression prediction models in whole blood, identifies significantly more gene-trait associations, and improves predictive power for identifying “silver standard” genes compared to several benchmark methods.

## 1 Introduction

Genome-wide association studies (GWASs) have identified large lists of disease-associated variants [1]. Most of the identified associations reside in non-coding regions of the genome [2], which has spurred the development of large-scale expression quantitative trait loci (eQTL) analyses [3–5] and transcriptome-wide association studies (TWASs) [6–12]. TWASs seek to investigate the causal molecular mechanisms underlying complex diseases through the integration of expression reference panels and trait-specific GWASs in a two-step procedure. First, a regression of gene expression on cis-eQTL genotypes is conducted to create expression prediction models for each gene. Second, the relationship between predicted gene expressions and GWAS traits are determined through an association test, elucidating putative causal effects of genes on traits of interest.

The power of TWAS is determined by the accuracy of the expression prediction model in Step 1 and the sample size of GWAS in Step 2 [6, 13]. As the sample size of GWAS continues to increase, thanks to the extensive consortium efforts, the prediction accuracy of the expression prediction model remains a limiting factor. To address this challenge, several methods have been proposed to improve the prediction model accuracy by leveraging auxiliary information from other tissues [10, 12], functional annotations or atlases of regulatory elements [14, 15], and summary-level expression reference panels (SUMMIT) [6]. As an example, by using the summary-level expression reference panel with a much larger sample size, SUMMIT outperforms many benchmark methods in terms of expression prediction model accuracy and subsequent TWAS power [6]. However, SUMMIT only relies on the summary-level expression reference panel and ignores comprehensive functional annotations that may be useful for improving expression prediction model accuracy. Functional annotations [16–18] assess functional roles for both coding and non-coding variants, providing a useful prior probability that a genetic variant causally affects the expression levels. We hypothesize that the accuracy of SUMMIT models (which are constructed by using summary-level expression reference panels) can be further improved by leveraging functional annotations, leading to higher power of TWAS.

To test our hypothesis, we develop SUMMIT-FA (Summary-level Unified Method for Modeling Integrated Transcriptome using Functional Annotations), an extension of the SUMMIT [6], that improves the accuracy of gene expression prediction models by leveraging annotation resources from the Multi-dimensional Annotation-Class Integrative Estimation (MACIE) database [18] to prioritize functional variants. MACIE synthesizes multiple categories of annotations, such as evolutionary conservation annotations and epigenetic annotations, and consistently provides the state-of-the-art performance in discriminating between functional and non-functional variants [18]. Briefly, we craft gene expression prediction models for whole-blood tissue using a summary-level eQTL database provided by the eQTLGen consortium [5], the largest-to-date publicly available meta-analysis featuring samples from 37 cohorts and 31,684 blood samples, and functional annotations provided by MACIE [18]. These models are then combined with the previous set of SUMMIT [6] models to significantly improve its performance. The elevated utility of SUMMIT-FA is thoroughly demonstrated via simulation studies and application to GWAS summary statistics for 24 complex traits. Notably, SUMMIT-FA increases expression prediction accuracy in whole-blood tissue, including for genes with low expression heritability, enhances power to detect associations between genes and phenotypes beyond preceding benchmark methods, and achieves higher predictive power for identifying “silver standard” genes, where “silver standard” genes are curated from information independent of GWAS results (see Methods section). A database of the SUMMIT-FA models and results is available as a resource to the community.

## 2 Results

### 2.1 SUMMIT-FA overview

SUMMIT-FA extends SUMMIT [6], a recently developed TWAS framework that leverages large-scale eQTL summary-level data to predict gene expression levels, to further improve accuracy in expression prediction and subsequent power for association testing. Briefly, SUMMIT-FA follows much the same scaffolding as SUMMIT: for each prospective gene, nine expression prediction models are trained on eQTL summary-level data from 31,684 whole-blood sample provided by eQTLGen [5]. Then, for each model with satisfactory imputation accuracy (*R*^2^ *>* 0.005), associations between predicted gene expression levels and phenotypes are tested. Given the potential correlation of *p*-values from the distinct models, the Cauchy combination test [19, 20] is applied to effectively and efficiently aggregate the nine separate results into a single one. The major advantage of SUMMIT-FA over its predecessor lies in four new gene expression prediction models that rely on MACIE [18] functional annotations. In short, a variant with a high MACIE_anyclass score (the posterior probability a variant is in some way functional, see [18] for details) will receive a lower penalty in the regression, resulting in prioritization of those variants believed to be more likely causal a priori. Additionally, each of the four models uses only SNPs with MACIE_anyclass greater than a given cutoff. For more details, see the Methods section.

### 2.2 SUMMIT-FA constructs more analyzable expression models

To demonstrate the elevated utility of SUMMIT-FA, we compared the model accuracy for gene expression prediction in whole blood tissue for SUMMIT-FA, SUMMIT (its predecessor) [6], and five benchmark methods: PrediXcan [8], TWAS-FUSION [7], UTMOST [10], MR-JTI [12], and Lassosum [21]. In an effort to make comparisons fair, we focused only on genes with estimated *R*^2^ ≥ 0.01 for SUMMIT-FA and SUMMIT (to match the other methods) that exist in eQTLGen data. PrediXcan, TWAS-FUSION, UTMOST, and MR-JTI were trained using GTEx data, and the prediction accuracies (*R*^2^) were based on a cross-validation procedure and provided by the authors. SUMMIT-FA, SUMMIT, and Lassosum were trained using summary-level eQTL data from the eQTLGen, and prediction accuracies were determined in an independent test dataset drawn from GTEx version 8 that was not included in the eQTLGen meta-analysis. Notably, SUMMIT-FA constructed satisfactory (in this case, *R*^2^ *>* 0.01) expression prediction models for more genes than the preceding methods: 12, 132 for SUMMIT-FA vs. 9, 749 for SUMMIT, 7, 512 for PrediXcan, 5, 411 for TWAS-FUSION, 7, 236 for UTMOST, 9, 576 for MR-JTI, and 8, 249 for Lassosum. Crucially, SUMMIT-FA successfully developed models for the majority of genes that SUMMIT and the benchmark methods did, 10, 018 (81.9%) out of the 12, 230 (unique genes across all other methods). Furthermore, SUMMIT-FA built satisfactory prediction models (with *R*^2^ ≥ 0.01) for an additional 950 genes beyond what was accomplished by SUMMIT or any of the five benchmark methods. Compared with SUMMIT alone, SUMMIT-FA built satisfactory prediction models (with *R*^2^ *>* 0.01) for additional 2,383 genes (24.4% improvement). These improvements demonstrate the marked utility of MACIE functional annotations in TWAS methods. The direct impact of their inclusion is seen as SUMMIT-FA achieved significantly higher prediction accuracy across the eQTLGen gene set than SUMMIT (*p <* 2.2 × 10^−16^ per the paired Wilcoxon rank-sum test) and other competing methods (PrediXcan, TWAS-FUSION, UTMOST, MR-JTI, and Lassosum; all *p <* 2.2 × 10^−16^ per the paired Wilcoxon rank-sum test).

### 2.3 SUMMIT-FA pinpoints significantly more associations

We explored the downstream performance of SUMMIT-FA in identifying significant associations by applying it to GWAS summary statistics of 24 complex phenotypes (see Supplementary Table 1 for a list of phenotypes, Supplementary Data 1 for full SUMMIT-FA association scan results). We then compared results from SUMMIT-FA to SUMMIT and the five aforementioned benchmark methods (PrediXcan, TWAS-FUSION, UTMOST, MR-JTI, and Lassosum). While both SUMMIT-FA and SUMMIT successfully analyze all genes with genetic heritability (i.e. prediction *R*^2^) *>* 0.5%, we first focused on all genes with heritability *>* 1% for fair comparison with other benchmark methods (Figure 2b). Across all traits, SUMMIT-FA identified 4,049 significant gene-phenotype associations, which is a 21.3% increase over SUMMIT (*p* = 7.2 × 10^−5^ via the one-sided Wilcoxon signed-rank test), a 161.4% increase over PrediXcan (*p* = 4.8 × 10^−5^), a 183.7% increase over TWAS-FUSION (*p* = 4.7 × 10^−5^), a 124.2% increase over UTMOST (*p* = 5.6 × 10^−5^), a 132.3% increase over MR-JTI (*p* = 9.5 × 10^−7^), and a 102.6% increase over Lassosum (*p* = 4.7 × 10^−5^).

**Figure 1:**
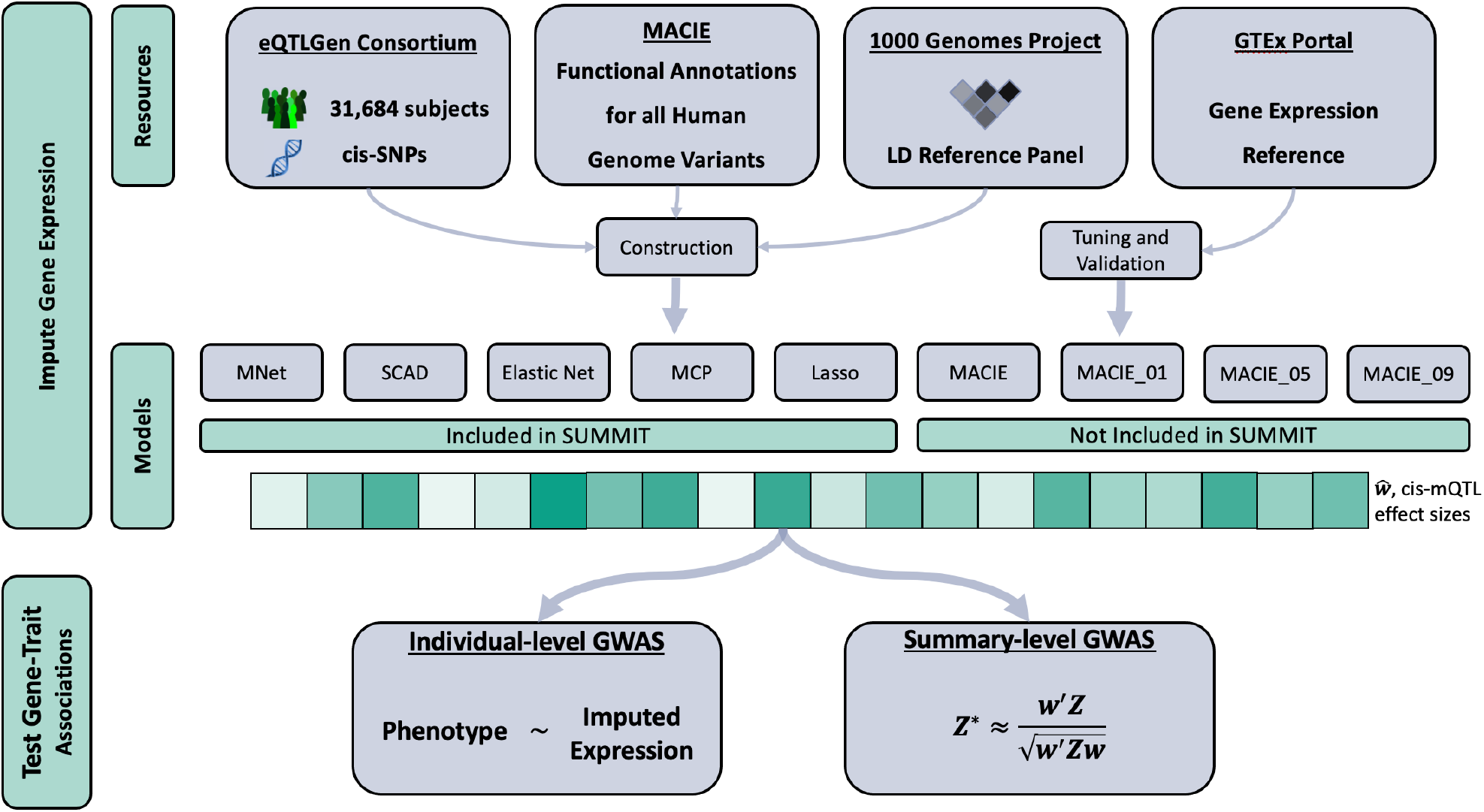
SUMMIT-FA Workflow. SUMMIT-FA proceeds in two steps. First, build gene expression prediction models. Second, test associations between traits of interest and imputed expression, aggregating the results from nine distinct prediction models.

**Figure 2:**
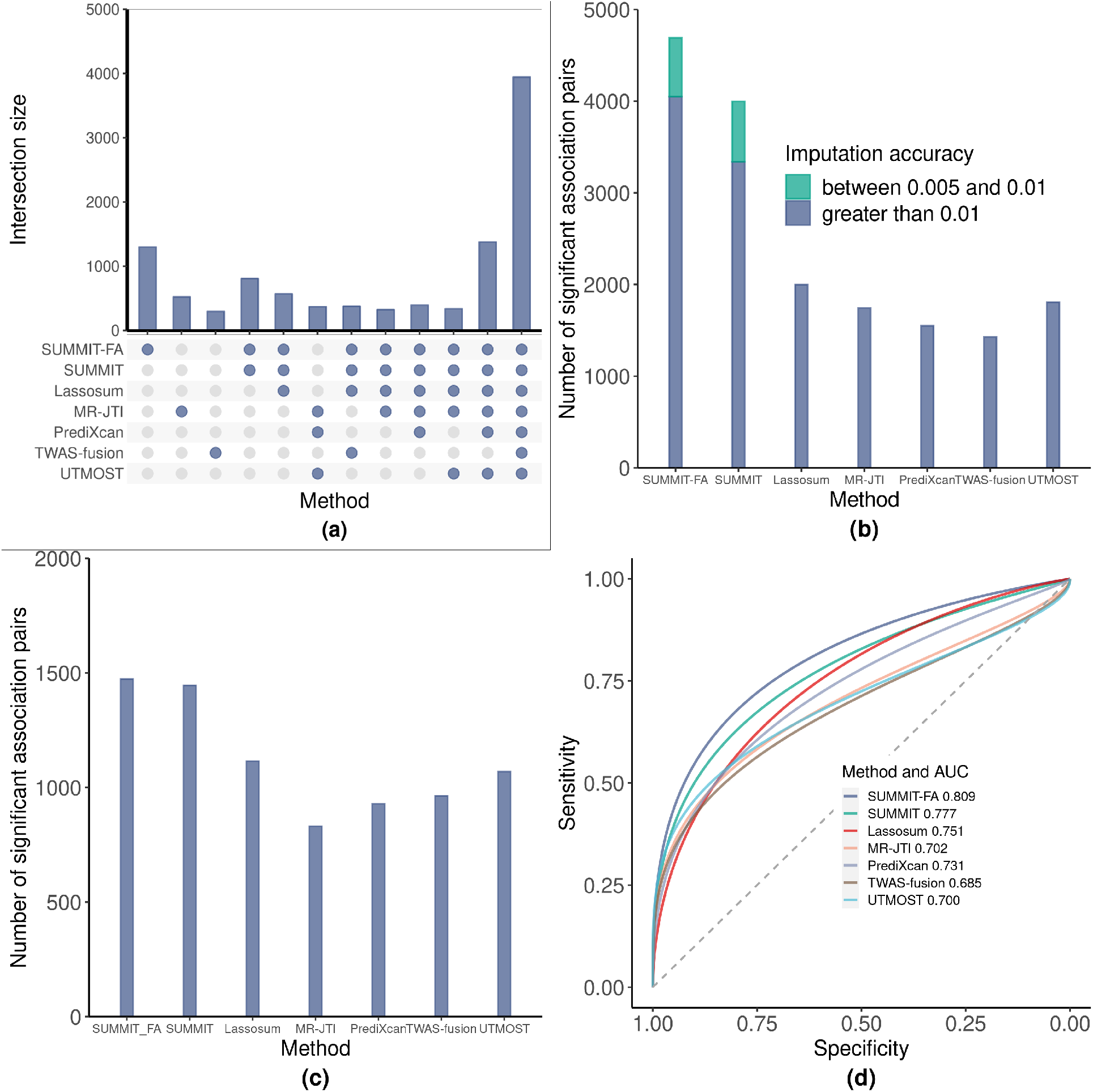
Method Comparison. a) UpSet plot on common gene set for which *R*^2^ ≥ 0.01. b) Number of significant gene-trait associations for all methods and genes across 24 GWASs. c) Number of significant gene-trait associations for all methods on a common set of genes across 24 GWASs. d) ROC plot for identifying silver standard genes.

We further compared each method on a common set of 3, 948 genes that could be analyzed by each (Figure 2c). For the common set of 3, 948 genes, both SUMMIT and SUMMIT-FA built expression prediction models with high prediction accuracy. The average *R*^2^ values for SUMMIT and SUMMIT-FA were 0.161 and 0.167, respectively. As a result, SUMMIT-FA and SUMMIT performed similarly and identified 1, 473 and 1, 445 significant associations, respectively, though the improvement of SUMMIT-FA over other benchmark methods is still quite large (a 32% increase in significant associations over the next highest-performing method, Lassosum). This result is aligned with our simulation study: including MACIE functional annotations is quite impactful for genes with lower expression heritability but significantly less so for genes with high expression heritability.

Lastly, recalling that SUMMIT provided a step forward in the field given the ability to analyze genes with lower heritability (0.005 ≤ *R*^2^ *<* 0.01), we examined these genes in more detail for SUMMIT-FA. For SUMMIT-FA, 14, 309 genes possess *R*^2^ ≥ 0.005; of these, 2, 177 have 0.005 ≤ *R*^2^ *<* 0.01. The 2, 177 lower heritability genes produce 642 significant gene-trait associations, a relatively similar ratio to that of genes with *R*^2^ ≥ 0.01: 12, 132 genes and 4, 049 significant genetrait associations. This gives further credence to the notion that lower heritability genes have notably larger causal effect sizes on complex phenotypes [6, 22].

### 2.4 SUMMIT-FA improves predictive power for identifying “silver standard” genes

We applied SUMMIT-FA and the benchmark methods to a set of “silver standard” genes, which are curated from information independent of GWAS results and considered highly likely to be causal in mediating associations between phenotypes and GWAS loci. As per Barbeira et al. [23], the silver standard gene set consists of 1, 258 gene-phenotype pairs from the Online Mendelian Inheritance in Man (OMIM) database [24] and an additional 29 pairs crafted from rare-variant results in exome-wide association studies. All silver standard gene-phenotype pairs are in Supplementary Data 2.

We used the silver standard gene set to compare sensitivity and specificity among the methods (Figure 2d). Notably, SUMMIT-FA produced the highest AUC (0.809), though the six benchmark methods (AUCs ∈ [0.685, 0.777]) still performed reasonably well, reinforcing the notion that TWAS methods can successfully identify putative causal genes. We further conducted a one-sided Delong test for the difference in AUC for the receiver-operator curves, resulting in a significant difference between SUMMIT-FA and TWAS-FUSION (*p* = 0.015), MR-JTI (*p* = 0.012), and UTMOST (*p* = 0.002). Potentially significant differences were found between SUMMIT-FA and SUMMIT (*p* = 0.056), PrediXcan (*p* = 0.080), and Lassosum (*p* = 0.058). Notably, the TWAS framework, including SUMMIT-FA, can be considered a type of Mendelian randomization or instrumental variable regression [25, 26]. The optimal instrument in this context is the prediction model with the highest accuracy [27]. As SUMMIT-FA increased the accuracy of its gene expression prediction models, it outperformed its contemporaries in identifying “silver standard” genes.

### 2.5 Simulation results

We conducted a simulation study to evaluate the accuracy and subsequent downstream power to detect associations of models produced by SUMMIT-FA using the gene *CHURC1*. We compared with its predecessor SUMMIT and three additional benchmark methods: Lassosum, TWAS-FUSION, and PrediXcan. We began by examining the accuracy, or *R*^2^, of gene expression prediction models built from the five methods (Figure 3a). SUMMIT-FA drastically outperforms the two methods that rely on individual-level expression reference panels (and thus have a significantly smaller sample size), TWAS-FUSION and PrediXcan. This is expected, as increasing the sample size of the reference has been shown to increase accuracy in gene expression imputation [6]. The two other methods that rely on larger, summary-level reference panels, SUMMIT and Lassosum, are significantly more accurate, though SUMMIT-FA still outperforms both. As an example, when 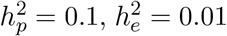, and *p*_causal_ = 0.05, the median prediction *R*^2^ for 1, 000 replicates for SUMMIT-FA is 0.81%, higher than the 0.75% for SUMMIT and 0.61% for Lassosum. We next demonstrated the elevated power of the downstream association tests for SUMMIT-FA (Figure 2b). Across different values for sparsity level (*p*_causal_) and expression heritability 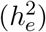, only SUMMIT approaches SUMMIT-FA’s power. For example, when 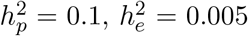, and *p*_causal_ = 0.05, the power for SUMMIT-FA is 0.798, 0.723 for SUMMIT, 0.282 for Lassosum, 0.002 for TWAS-FUSION, and 0.0 for PrediXcan.

**Figure 3:**
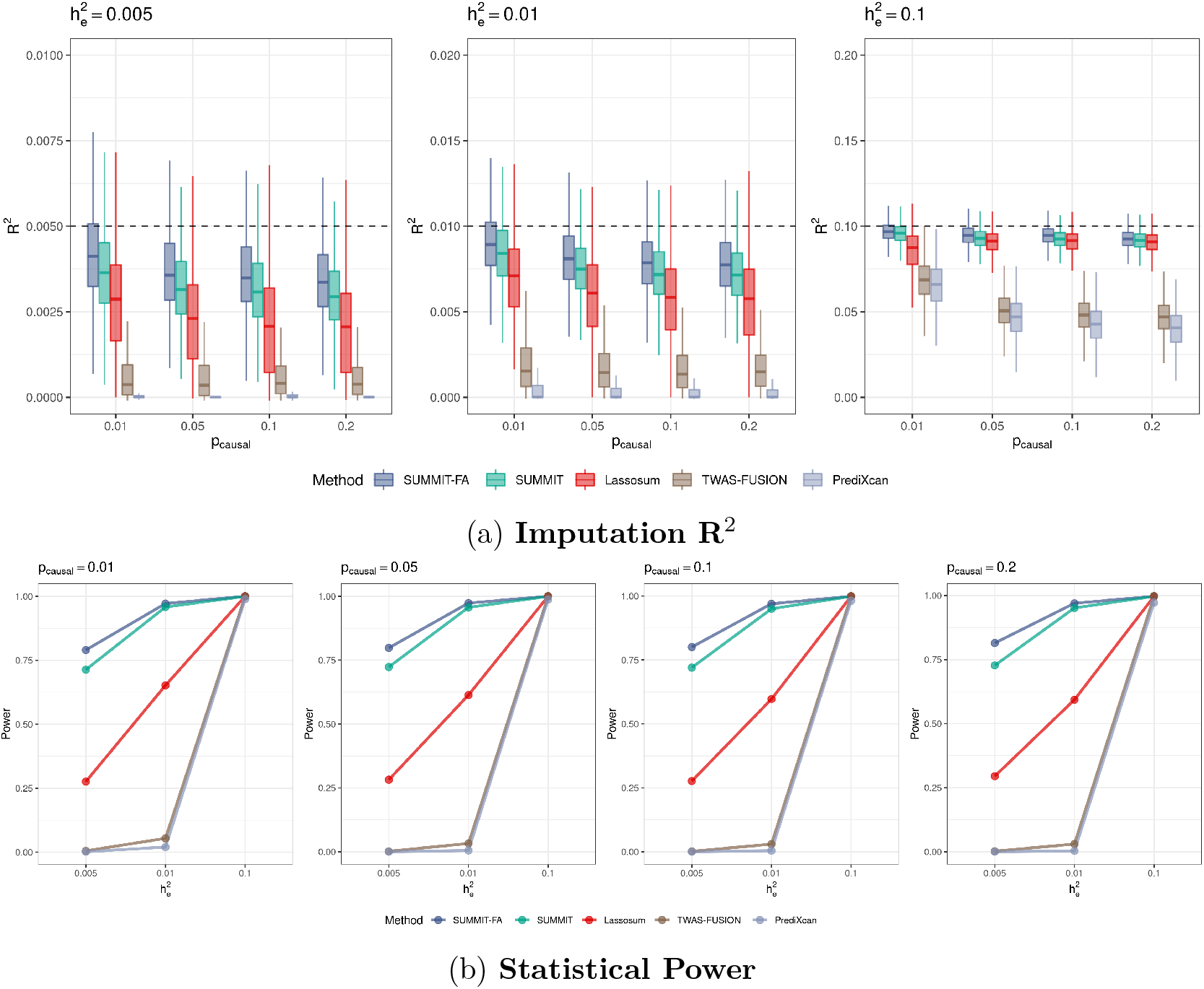
Simulation performance comparison based on gene CHURC1. a) Plots of imputation *R*^2^ in test samples by SUMMIT-FA, SUMMIT, Lassosum, TWAS-FUSION, and PrediXcan for varied proportion of causal SNPs p_causal_ and DNAm heritability 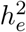, with phenotypic heritability 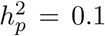. b) Subsequent power measurements for each method with phenotypic heritability 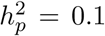. Empirical power was determined by the proportion of p-values *<* 2.5 × 10^−6^.

It has been shown that genes with lower expression heritability (i.e. prediction *R*^2^ ∈ [0.005, 0.01)) generally have larger effect sizes on complex phenotypes [22] and thus are important to be tested in TWAS [6]. As such, it is paramount that SUMMIT-FA retains SUMMIT’s ability to achieve satisfactory performance for genes with low expression heritability. We simulated data with 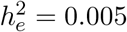 to examine this, which is shown in Figure 3a. SUMMIT-FA consistently achieves satisfactory results; as an example, when 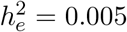, and *p*_causal_ = 0.05, SUMMIT-FA’s median prediction *R*^2^ is 0.36%, 14.6% higher than SUMMIT’s 0.31%.

We demonstrated consistent results for an additional randomly selected gene, *KRIT1*, to ensure that genetic architecture is not the driving force of the simulation results (Supplementary Figures 4-5). We further ran 20, 000 simulation replicates under the null hypothesis to evaluate Type 1 error rates. Every method controlled Type 1 error rates well (Supplementary Figure 3). To summarize, the simulation results demonstrate the viability of SUMMIT-FA models for use in predicting gene expression and subsequent downstream association tests. Importantly, SUMMIT-FA’s elevated performance beyond its predecessor SUMMIT, in particular in the low expression heritability setting, demonstrated the advantages of including MACIE functional annotations in the model construction.

## 3 Discussion

By including functional annotations provided by MACIE [18], models built by the new method SUMMIT-FA achieve increased gene expression prediction accuracy beyond those of its predecessor SUMMIT [6], which subsequently increased the downstream power to detect risk genes for complex phenotypes. Given its performance in the analyses of 24 distinct GWAS and a simulation study, SUMMIT-FA marks a substantial step forward from preceding methods. Specifically, SUMMIT-FA improved gene expression prediction accuracy, pinpointed more gene-trait associations, was better powered to detect silver standard genes, and, importantly, maintained and improved upon its predecessor’s ability to analyze genes with low heritability of expression (i.e. *R*^2^ ∈ [0.005, 0.01)). These genes generally have larger effect sizes on complex phenotypes [6, 22], and as such new TWAS methods ought to ensure they remain analyzable.

As a TWAS method, SUMMIT-FA may be viewed as one type of Mendelian randomization (MR) [25, 26]. Thus, it can allow for valid causal interpretations, but only when every genetic variant included in the gene expression prediction models is a valid instrument variable (IV) [26, 28, 29]. Given that horizontal pleiotropy is widespread [30], it is likely that IV assumptions are violated, and we in turn recommend that those who wish to make use of this resource do so along with other complementary methods, such as fine-mapping [31, 32] or colocalization [33], both of which can prioritize putatively causal genes in different aspects.

Naturally, SUMMIT-FA is largely motivated by our previous method SUMMIT [6] and the recent development of the MACIE functional annotations [18], which, unlike preceding annotation sets, cover the entirety of the genome. While some TWAS methods integrate additional datasets beyond GWASs and gene expression panels [10, 12, 34], so far as we are aware, none incorporate comprehensive functional annotation databases like MACIE (which has been shown to outperform other such annotations [18]) in the same manner as SUMMIT-FA. As demonstrated in this work, making use of MACIE can improve overall TWAS performance.

We conclude with a discussion of limitations for the present study. Primarily, the summary-level expression reference from eQTLGen comes from subjects of European ancestry in whole blood tissue, and thus SUMMIT-FA models (as presently built) can only be applied successfully to such data. In theory, SUMMIT-FA can be applied to summary-level eQTL data from any tissue and ancestry, but these datasets do not yet exist in any appreciable size. Additionally, we focused on *cis*-eQTLs in the gene expression prediction models and did not consider *trans*-eQTLs. Furthermore, as demonstrated in one recent study [15], 3D genomic and epigenomic data provide additional information and can be used to further improve the accuracy of expression prediction models. We expect that incorporating *trans*-acting elements or leveraging auxilary information from other tissues, ancestry groups, and 3D genomic and epigenomic can further improve performance, though substantial additional work would be necessary. Lastly, as discussed above, causal arguments based on SUMMIT-FA (as with any TWAS method) bear scrutiny only when IV assumptions are not violated. Thus, caution and complementary methodology are suggested with regard to arguing causality. We leave these exciting topics for future studies.

## 4 Methods

### 4.1 Predicting gene expression with penalized regression models

We create nine gene expression predictions models through penalized regression. Begin with the following model:

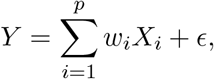

where *Y* is the *N* -dimensional vector of gene expression levels for a particular gene corrected for age, sex, and genetic principal components, 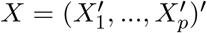 is the standardized genotype matrix of *cis*-SNPs around the gene, *w* = (*w*_1_, …, *w*_*p*_)^′^ is the eQTL effect size (i.e. the weights), and *ϵ* is mean-zero random noise. We then estimate *w* with a penalized regression framework.

SUMMIT-FA inherits many of the advantages of SUMMIT, including five gene expression prediction models with distinct penalties, stability from using a shrinkage estimator of the LD matrix, and the ability to combine results across models by the use of the Cauchy combination test [20]. Briefly, the five inherited models estimate eQTL effect sizes *w* by optimizing the objective function:

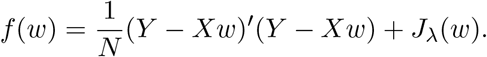

Through simplification and recognizing that 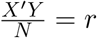, the standardized marginal effect sizes for the *cis*-SNPs (the correlation between gene expression levels and these SNPs), and 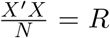, the LD matrix, we arrive at the following objective function:

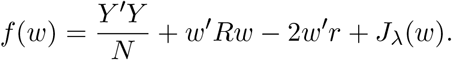

Following the SUMMIT [6], we then estimate the LD matrix *R* by a shrinkage estimator of a reference panel, such as from the 1000 Genomes project [35] denoted 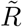, and we estimate the standardized marginal effects *r* with 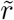, which come from the z-scores provided by the summary-level statistics in the eQTLGen database. We note that 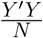 is not a function of *w* and can thus be safely ignored in the optimization procedure. Thus, the final objective function is:

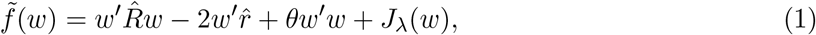

where *J*_*λ*_(·) is the main penalty term. Five penalties are employed here, LASSO [36], elastic net [37], the minimax concave penalty (MCP) [38], the smoothly clipped absolute deviation (SCAD) [39], and MNet [40]. *θw*^′^*w, θ* ≥ 0 is an additional *L*_2_ penalty designed to ensure a unique solution for 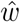. We then optimize 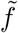 via the coordinate descent algorithm [41], which is further discussed in the following section.

#### 4.1.1 Incorporating MACIE functional annotations into penalized regression framework

MACIE (Multi-dimensional Annotation-Class Integrative Estimation) [18] annotations are a vector of length four of joint posterior functional probabilities of functional roles for variants. MACIE_anyclass is the sum of these four probabilities, and represents a prior (with respect to SUMMIT-FA’s gene expression prediction models) estimate of the functional probability according to at least one class of annotations of variants; a higher value indicates a higer chance to be functional. We incorporate MACIE_anyclass in two distinct ways to the model. First, we restrict the eQTL set to include variants with a likelihood greater than some threshold *c* of functional behavior, so the variant set becomes {*X*_*i*_ : MACIE_anyclass_*i*_ ≥ *c*}, with *c* ∈ {0.0, 0.1, 0.5, 0.9}. The four thresholds lead to the four new models in SUMMIT-FA, each of which uses an elastic net penalized regression.

Second, we employ MACIE_anyclass in the elastic net penalty. From equation (1), we define the penalty for SNP 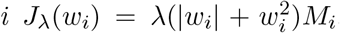, which is the typical elastic net penalty with *α* = 0.5 multiplied by *M*_*i*_, where *M*_*i*_ is one minus MACIE_anyclass value for SNP *i*. Thus, we reduce the penalty for SNPs that have, a priori, a higher likelihood of functionality. These SNPs are considered more likely to be causal, and are therefore included in the gene expression models with greater frequency under this method.

The solution to the optimization, *ŵ*, is then constructed via the coordinate descent algorithm [41]. Denote by 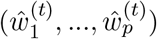 the t-th realization of the coordinate descent process, and let 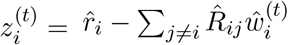. Then the (*t* + 1)th update of *ŵ*_*i*_ is

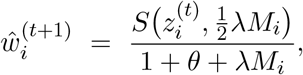

for *i* = 1, …, *p* and *t* = 0, 1, 2, …, where *S*(*V, ω*) is the soft-thresholding operator

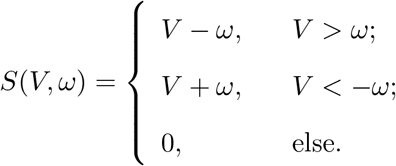

The coordinate descent algorithm then converges to a local minimum for *ŵ* [41]. Tuning parameters *θ*, which is restricted to the set {0.1, 0.2, …, 0.9}, and *λ*, which has a solution path generated by the warm start algorithm, are chosen to maximize *R*^2^ in the training data.

### 4.2 Training and Evaluating Model

Gene expression predictions models were trained on eQTL summary data from eQTLGen [5], which includes effect sizes of more than 11 million SNPs derived from 31, 684 blood samples. We employed only *cis*-SNPs, those within 1 Mbp of gene transcription start and end sites. We further filtered out any SNPs that were nonbiallelic, ambiguous, not included in the HapMap3 SNP set [8], or had minor allele frequency (MAF) *<* 0.01.

To choose tuning parameters, we used genotype and gene expression references from the GTEx project (version V7, dbGaP Accession number phs000424.v7.p2, https://www.gtexportal.org/home/datasets) [42]. The whole blood gene expression levels (*N* = 369) were processed by standardizing and normalizing RPKMs in each sample then adjusting for sex, genotyping platform, 35 PEER factors and three genotype-based principal components. Residuals were then used as the processed gene expression levels, which downloaded from the GTEX website. We then selected optimal tuning parameters based on the *R*^2^ (i.e. squared correlation between predicted and observed expression levels. We note that subjects in GTEx v6 (*N* = 336; 1.1%) were included in eQTLGen’s meta analysis [5], which may result in suboptimal tuning decisions.

We conducted an external validation with a fully independent set of subjects (*N* = 309) in GTEx v8 that were not included in GTEx v7 nor in the eQTLGen meta-analysis. Following SUMMIT [6], genes with estimated expression heritability (*R*^2^) of at least 0.5% in the validation dataset were included in downstream association analysis, as genes with low heritability have larger causal effect sizes on complex traits [22].

### 4.3 Conducting Association Analyses

With individual-level GWAS data, we employ a generalized linear regression

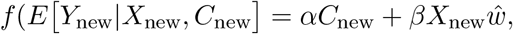

where *X*_new_ is the genotype data, *Y*_new_ is the phenotype data, and *C*_new_ is the covariant matrix, *f* is a link function, and *X*_new_*ŵ* is the predicted gene expression levels. We then test *H*_0_ : *β* = 0 for gene-trait associations.

With summary-level GWAS data, we apply a burden-style test

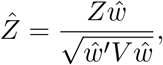

where *Z* is a vector of z-scores for all *cis*-SNPs and *V* is the LD matrix for all SNPs included in the analysis. *V* is typically estimated with an LD reference panel (such as the 1000 Genomes Project [35]).

Note that the above descriptions for conducting association analysis assumes only a single expression prediction model, but SUMMIT-FA includes up to nine distinct models. In the case of multiple methods building satisfactory models at a gene (i.e. gene expression prediction *R*^2^ *>* 0.005 for that gene), each follows the aforementioned procedure to conduct an association test. Then, results are amalgamated on the gene-level via the Cauchy combination test [30], which has been widely used in the field [6, 34]. Briefly, assume *K* satisfactory models and consider the test statistic:

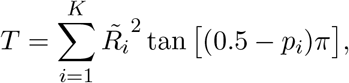

where 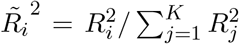 and *p*_*i*_ is the p-value for the *i*th model. *T* approximately follows the standard Cauchy distribution, and the overall p-value is calculated by *p* = 0.5 − arctan(*T*)*/π*. We note that the test statistic is a sum weighted by 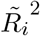, which follows logically from the notion that larger values of 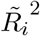 come from better fitting gene expression prediction models.

### 4.4 Simulation setup

We demonstrated the elevated performance SUMMIT-FA through a set of comprehensive simulation studies. Both gene expression prediction accuracy and downstream TWAS power were evaluated, and, additionally, SUMMIT-FA was shown to successfully build gene expression prediction models using summary-level eQTL data. We used genotype data from 31, 684 (to was shown match the sample size of the eQTLGen data) white British subjects from the UK Biobank as training data, genotype data from 369 (to match the sample size of the GTEx v7 data) independent white British subjects from the UK Biobank as tuning data, and genotype data from 10, 000 more independent white British subjects from the UK Biobank as testing data. Imputed data from 877 *cis*-SNPs (with MAF *>* 1%, Hardy-Weinberg *p*-value *>* 10^−6^, and imputation “info” score *>* 0.4) of the randomly selected gene *CHURC1* were in primary simulations. Another randomly selected gene *KRIT1* is considered in Supplementary Figures 3, 4.

We compare SUMMIT-FA to SUMMIT [6], PrediXcan [8], TWAS-fusion [7], and Lassosum [21] in terms of gene expression prediction accuracy and downstream power for TWAS. Simulation settings varied the phenotypic heritability 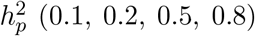, expression heritability 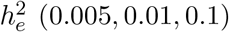, and proportion of causal SNPs *p*_causal_ (0.01, 0.05, 0.1, 0.2). We considered scenarios that varied the proportion of causal SNPs *p*_causal_ (0.01, 0.05, 0.1, 0.2), expression heritability 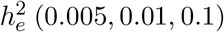, and phenotypic heritability 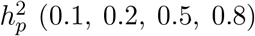. For each scenario, we repeated the simulations 1,000 times. The statistical power was determined by the proportion of 1,000 repeated simulations with association *p*-value less than the genome-wide significance threshold 0.05*/*20, 000 = 2.5 × 10^−6^.

Gene expression levels are simulated according to *E*_*g*_ = *Xw* + *ϵ*_*e*_, and phenotypes by *Y* = *βE*_*g*_ + *ϵ*_*p*_, where *X* is the standardized genotype matrix, *w* the effect size, *E*_*g*_ gene expression levels, *β* the association coefficient, 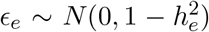, and 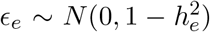. 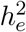 is the expression heritability, the proportion of variance in gene expression explained by SNPs, and 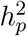 is the phenotype heritability, the proportion of variance in gene expression explained by gene expression. Given *M* SNPs potentially included in the model, *l* are selected to have nonzero effect size in order to achieve the desired *p*_causal_. The *l* SNPs are sampled from the *M* total SNPs with probabilities proportional to their MACIE_anyclass scores. The effect sizes *w* and *β* were then rescaled to achieve the correct values for 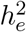 and 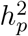.

With regard to the other methods involved in the simulation, PrediXcan and TWAS-fusion were trained on individual-level genotype data from 670 white British samples (matching the sample size of blood tissue in the GTEx v8 data). Lassosum and SUMMIT, which can employ summary-level data, were trained on the same summary-level data as that used for SUMMIT-FA. All models were compared on single-tissue eQTL information, as cross-tissue information was not our focus here. As such, we chose not to include UTMOST [10] and MR-JTI [12], as it would be disingenuous to compare them without making use of further tissues, the main contribution of these two methods. This topic is left to further research.

### 4.5 Comparison with existing methods

To further demonstrate the added utility provided by SUMMIT-FA, we compared it to six previous TWAS methods in whole blood tissue: SUMMIT [6], PrediXcan [8], TWAS-fusion [7], UTMOST [10], MR-JTI [12], and Lassosum [21]. To briefly describe each, SUMMIT is the predecessor to SUMMIT-FA; it features five gene expression prediction models that are combined with the Cauchy combination test, though it does not include functional annotations. PrediXcan uses the Elastic Net to create expression prediction models. TWAS-Fusion applies BLUP, BSLMM, Elastic Net, LASSO, and TOP1 in building models for expression prediction. MR-JTI and UTMOST leverage cross-tissue eQTLs when building gene expression prediction models. Lastly, Lassosum is actually a method to create polygenic risk scores, though it can be used to build gene expression prediction models from summary-level eQTL reference panels, which are then applied to the TWAS setup. SUMMIT-FA, SUMMIT, and Lassosum utilize summary-level reference datasets, while the other four TWAS models require individual-level reference panels.

We begin by examining the gene expression prediction accuracy (i.e. *R*^2^) produced by the distinct methods. We note that while *R*^2^ is estimated in a test dataset for SUMMIT-FA, SUMMIT, and Lassosum, it is determined by cross-validation for the others, which may marginally favor PrediXcan, TWAS-Fusion, UTMOST, and MR-JTI. To demonstrate the utility of functional annotations provided by the MACIE, we tested the difference between estimated expression prediction accuracy for SUMMIT-FA and SUMMIT with a paired Wilcoxon rank-sum test, which is a nonparametric test that compares two matched samples to test whether population mean ranks differ.

We then applied the methods to analyses of 24 sets of GWAS summary statistics for complex phenotypes (the traits are detailed in Supplementary Data 1). We applied the Bonferroni correction to each method with distinct thresholds, as each method analyzed a different number of genes. In the interest of fairness, we also used a common set of genes for which all models could analyze and identical Bonferroni-generated significance cutoffs. We then applied a one-sided Wilcoxon signed-rank test to the numbers of significant genes identified by the different methods.

Lastly, we compared the methods ability to pinpoint causal genes that mediate associations between phenotypes and GWAS loci. We created a set of likely causal gene-trait pairs by following Barbeira et al. [23] that was independent of the GWAS results. We obtained 1, 287 gene-trait pairs using OMIM [24] and rare variant results from exome-wide association studies [43–45]. We employed LDetect to partition the genome into presumably independent LD blocks [46], and only included gene-trait pairs living in LD blocks with genome-wide significant variants. This resulted in 148 putatively causal pairs across 24 traits. We finally compared the methods by measuring the area under the receiver operator curve (ROC) and tested whether the area under the curve (AUC) differences were significant using a one-sided Delong test. Parametric bootstrap is also used for assessing the AUC differences and the results were similar (not shown).

## Data Availability

The GWAS summary data used in this study are summarized in Supplementary Data 1 (with the download link). The eQTL summary data are available at https://www.eqtlgen.org/cis-eqtls. html The UK Biobank is an open-access resource requiring registration, available at https://www.ukbiobank.ac.uk/researchers/. The genotype and RNA sequencing data for the GTEx project may be found at the database of Genotypes and Phenotypes (accession number phs000424.v8.p2, https://www.ncbi.nlm.nih.gov/projects/gap/cgi-bin/study.cgi?study_id=phs000424.v8.p2). The processed gene expression data from the GTEx project is available from the GTEx portal (https://gtexportal.org). The MR-JTI, PrediXcan, and UTMOST models may be downloaded from https://doi.org/10.5281/zenodo.3842289. The TWAS-FUSION models may be down-loaded from http://gusevlab.org/projects/fusion/. The 1000 Genomes Project data may be downloaded from https://www.internationalgenome.org/data. The genetic distance data for 1000 Genomes Project may be downloaded from https://github.com/joepickrell/1000-genomes-genetic-maps. The SUMMIT-FA models generated for this study will be available at OSF.IO.

https://www.eqtlgen.org/cis-eqtls.html

https://www.ukbiobank.ac.uk/researchers/

https://www.ncbi.nlm.nih.gov/projects/gap/cgi-bin/study.cgi?study_id=phs000424.v8.p2

https://gtexportal.org/

https://doi.org/10.5281/zenodo.3842289

http://gusevlab.org/projects/fusion/

https://www.internationalgenome.org/data

https://github.com/joepickrell/1000-genomes-genetic-maps

## Data availability

The GWAS summary data used in this study are summarized in Supplementary Data 1 (with the download link). The eQTL summary data are available at https://www.eqtlgen.org/cis-eqtls.html The UK Biobank is an open-access resource requiring registration, available at https://www.ukbiobank.ac.uk/researchers/. The genotype and RNA sequencing data for the GTEx project may be found at the database of Genotypes and Phenotypes (accession number phs000424.v8.p2, https://www.ncbi.nlm.nih.gov/projects/gap/cgi-bin/study.cgi?study_id=phs000424.v8.p2). The processed gene expression data from the GTEx project is available from the GTEx portal (https://gtexportal.org). The MR-JTI, PrediXcan, and UTMOST models may be downloaded from https://doi.org/10.5281/zenodo.3842289. The TWAS-FUSION models may be downloaded from http://gusevlab.org/projects/fusion/. The 1000 Genomes Project data may be downloaded from https://www.internationalgenome.org/data. The genetic distance data for 1000 Genomes Project may be downloaded from https://github.com/joepickrell/1000-genomes-genetic-maps. The SUMMIT-FA models generated for this study will be available at OSF.IO at. The raw data and code to replicate figures and tables in the manuscript will also be available at OSF.IO at.

## Code availability

Code to use the SUMMIT-FA modles will be available at GitHub (https://github.com/ChongWuLab/SUMMIT) and Zenodo. The codes and corresponding data for reproducing the results described in this study will be available on OSF.IO.

## Acknowledgements

National Institutes of Health (R03 AG070669) supported Z.Z., J.B., and C.W.. This study was conducted using the UK Biobank recourse under Application Number 48240 (https://www.ukbiobank.ac.uk/researchers/). The content is solely the responsibility of the authors and does not necessarily represent the official views of the National Institutes of Health. The Genotype-Tissue Expression (GTEx) Project was supported by the Common Fund of the Office of the Director of the National Institutes of Health, and by NCI, NHGRI, NHLBI, NIDA, NIMH, and NINDS. The authors would like to thank all of the individuals for their participation in the GWASs and UK Biobank and all the researchers, clinicians, technicians and administrative staff for their contribution to the studies and for making their GWAS summary results publicly available.

## Author contributions statement

C.W. conceived and designed the study. H.M and Z.Z developed the computational algorithms and wrote the SUMMIT-FA program. H.M performed the real data analysis and simulations. Z.Z. created the website that curated the results. H.M tested the program and drew the workflow diagram of SUMMIT-FA. All authors wrote and proofread the manuscript.

## References

[1] Annalisa Buniello et al. “The NHGRI-EBI GWAS Catalog of published genome-wide association studies, targeted arrays and summary statistics 2019”. In: Nucleic Acids Research 47 (D1 Jan. 2019), pp. D1005–D1012. ISSN: 0305-1048.

[2] Matthew T. Maurano et al. “Systematic localization of common disease-associated variation in regulatory DNA”. In: Science 337 (6099 Sept. 2012), pp. 1190–1195. ISSN: 10959203.

[3] GTEx Consortium. “The GTEx Consortium atlas of genetic regulatory effects across human tissues”. In: Science 369 (6509 2020), pp. 1318–1330.

[4] Yi Yang, Kar Fu Yeung, and Jin Liu. “CoMM-S4: A Collaborative Mixed Model Using Summary-Level eQTL and GWAS Datasets in Transcriptome-Wide Association Studies”. In: Frontiers in Genetics 12 (Sept. 2021), p. 1820. ISSN: 16648021.

[5] Urmo VÃsa et al. “Large-scale cis- and trans-eQTL analyses identify thousands of genetic loci and polygenic scores that regulate blood gene expression”. In: Nature Genetics 2021 53:9> 53 (9 Sept. 2021), pp. 1300–1310. ISSN: 1546-1718.

[6] Zichen Zhang et al. “SUMMIT: An integrative approach for better transcriptomic data imputation improves causal gene identification”. In: Nature Communications 2022 13:1> 13 (1> Oct. 2022), pp. 1–12. ISSN: 2041-1723.

[7] Alexander Gusev et al. “Integrative approaches for large-scale transcriptome-wide association studies”. In: Nature Genetics (2016). ISSN: 15461718.

[8] Eric R. Gamazon et al. “A gene-based association method for mapping traits using reference transcriptome data”. In: Nature Genetics 47 (9> 2015), pp. 1091–1098. ISSN: 15461718>.

[9] Zhiyuan Xu et al. “A powerful framework for integrating eqtl and gwas summary data”. In: Genetics 207 (3Nov. 2017), pp. 893–902. ISSN: 19432631.

[10] Yiming Hu et al. “A statistical framework for cross-tissue transcriptome-wide association analysis”. In: Nature Genetics 2019 51:3> 51 (3Feb. 2019), pp. 568–576. ISSN: 1546-1718.

[11] Sini Nagpal et al. “TIGAR: An Improved Bayesian Tool for Transcriptomic Data Imputation Enhances Gene Mapping of Complex Traits”. In: American Journal of Human Genetics 105 (2Aug. 2019), pp. 258–266. ISSN: 15376605.

[12] Dan Zhou et al. “A unified framework for joint-tissue transcriptome-wide association and Mendelian randomization analysis”. In: Nature Genetics 2020 52:11> 52 (11Oct. 2020), pp. 1239–1246. ISSN: 1546-1718.

[13] Ruoyu He, Haoran Xue, and Wei Pan. “Statistical power of transcriptome-wide association studies”. In: Genetic Epidemiology 46 (8Dec. 2022), pp. 572–588. ISSN: 1098-2272.

[14] Wen Zhang et al. “Integrative transcriptome imputation reveals tissue-specific and shared biological mechanisms mediating susceptibility to complex traits”. In: Nature Communications 2019 10:1 10 (1Aug. 2019), pp. 1–13. ISSN: 2041-1723.

[15] Chachrit Khunsriraksakul et al. “Integrating 3D genomic and epigenomic data to enhance target gene discovery and drug repurposing in transcriptome-wide association studies”. In: Nature Communications 2022 13:1> 13 (1June 2022), pp. 1–15. ISSN: 2041-1723.

[16] Mark F. Rogers et al. “FATHMM-XF: accurate prediction of pathogenic point mutations via extended features”. In: Bioinformatics 34 (3Feb. 2018), p. 511. ISSN: 14602059.

[17] Xihao Li et al. “Dynamic incorporation of multiple in silico functional annotations empowers rare variant association analysis of large whole-genome sequencing studies at scale”. In: Nature Genetics 52 (92020), pp. 969–983. ISSN: 15461718.

[18] Xihao Li et al. “A multi-dimensional integrative scoring framework for predicting functional variants in the human genome”. In: American journal of human genetics 109 (3Mar. 2022), pp. 446–456. ISSN: 1537-6605.

[19] Yaowu Liu and Jun Xie. “Cauchy Combination Test: A Powerful Test With Analytic p-Value Calculation Under Arbitrary Dependency Structures”. In: https://doi.org/10.1080/01621459.2018.1554485 115 (529Jan. 2019), pp. 393–402. ISSN: 1537274X.

[20] Yaowu Liu et al. “ACAT: A Fast and Powerful p Value Combination Method for Rare-Variant Analysis in Sequencing Studies”. In: American Journal of Human Genetics 104 (3Mar. 2019), p. 410. ISSN: 15376605.

[21] Timothy Shin Heng Mak et al. “Polygenic scores via penalized regression on summary statistics”. In: Genetic Epidemiology 41 (6Sept. 2017), pp. 469–480. ISSN: 1098-2272.

[22] Douglas W. Yao et al. “Quantifying genetic effects on disease mediated by assayed gene expression levels”. In: Nature Genetics 2020 52:6> 52 (6 May 2020), pp. 626–633. ISSN: 1546-1718.

[23] Alvaro N. Barbeira et al. “Exploiting the GTEx resources to decipher the mechanisms at GWAS loci”. In: Genome Biology 22 (1Dec. 2021), pp. 1–24. ISSN: 1474760X.

[24] Ada Hamosh et al. “Online Mendelian Inheritance in Man (OMIM), a knowledgebase of human genes and genetic disorders”. In: Nucleic Acids Research 33 (suppl_1 Jan. 2005), pp. D514–D517. ISSN: 0305-1048.

[25] Brandon L. Pierce and Stephen Burgess. “Efficient Design for Mendelian Randomization Studies: Subsample and 2-Sample Instrumental Variable Estimators”. In: American Journal of Epidemiology 178 (7Oct. 2013), pp. 1177–1184. ISSN: 0002-9262.

[26] Stephen Burgess and Simon G. Thompson. “Use of allele scores as instrumental variables for Mendelian randomization”. In: International Journal of Epidemiology 42 (4Aug. 2013), pp. 1134–1144. ISSN: 0300-5771.

[27] A. Belloni et al. “Sparse Models and Methods for Optimal Instruments With an Application to Eminent Domain”. In: Econometrica 80 (6Nov. 2012), pp. 2369–2429. ISSN: 1468-0262.

[28] Zhongshang Yuan et al. “Testing and controlling for horizontal pleiotropy with probabilistic Mendelian randomization in transcriptome-wide association studies”. In: Nature Communications 2020 11:1 11 (1July 2020), pp. 1–14. ISSN: 2041-1723.

[29] Haoran Xue and Wei Pan. “Some statistical consideration in transcriptome-wide association studies”. In: Genetic Epidemiology 44 (3Apr. 2020), pp. 221–232. ISSN: 1098-2272.

[30] Xuanyao Liu, Yang I. Li, and Jonathan K. Pritchard. “Trans Effects on Gene Expression Can Drive Omnigenic Inheritance”. In: Cell 177 (4May 2019), 1022–1034.e6. ISSN: 1097-4172.

[31] Nicholas Mancuso et al. “Probabilistic fine-mapping of transcriptome-wide association studies”. In: Nature Genetics (2019). ISSN: 15461718.

[32] Chong Wu and Wei Pan. “A powerful fine-mapping method for transcriptome-wide association studies”. In: Human genetics 139 (2Feb. 2020), p. 199. ISSN: 14321203.

[33] Claudia Giambartolomei et al. “Bayesian Test for Colocalisation between Pairs of Genetic Association Studies Using Summary Statistics”. In: PLOS Genetics 10 (52014), e1004383. ISSN: 1553-7404.

[34] Chong Wu et al. “A gene-level methylome-wide association analysis identifies novel Alzheimer’s disease genes”. In: Bioinformatics 37 (14July 2021), p. 1933. ISSN: 14602059.

[35] 1000 Genomes Project Consortium. “A global reference for human genetic variation”. In: Nature 2015 526:7571 526 (7571Sept. 2015), pp. 68–74. ISSN: 1476-4687.

[36] Robert Tibshirani. “Regression Shrinkage and Selection Via the Lasso”. In: Journal of the Royal Statistical Society: Series B (Methodological) 58 (1Jan. 1996), pp. 267–288. ISSN: 2517-6161.

[37] Hui Zou and Trevor Hastie. “Regularization and variable selection via the elastic net”. In: Journal of the Royal Statistical Society: Series B (Statistical Methodology) 67 (2Apr. 2005), pp. 301–320. ISSN: 1467-9868.

[38] Cun-Hui Zhang. “Nearly unbiased variable selection under minimax concave penalty”. In: The Annals of Statistics 38 (22010), pp. 894–942.

[39] Jianqing Fan and Runze Li. “Variable Selection via Nonconcave Penalized Likelihood and its Oracle Properties”. In: Journal of the American Statistical Association 96 (456Dec. 2001), pp. 1348–1360. ISSN: 0162-1459.

[40] Jian Huang et al. “The Mnet method for variable selection”. In: Statistica Sinica 26 (3July 2016), pp. 903–923. ISSN: 10170405.

[41] Jerome Friedman, Trevor Hastie, and Rob Tibshirani. “Regularization Paths for Generalized Linear Models via Coordinate Descent”. In: Journal of statistical software 33 (12010), p. 1. ISSN: 15487660.

[42] GTEx Consortium. “Genetic effects on gene expression across human tissues”. In: Nature 2017 550:7675 550 (7675Oct. 2017), pp. 204–213. ISSN: 1476-4687.

[43] Dajiang J. Liu et al. “Exome-wide association study of plasma lipids in >300,000 individuals”. In: Nature Genetics 2017 49:12 49 (12Oct. 2017), pp. 1758–1766. ISSN: 1546-1718.

[44] Eirini Marouli et al. “Rare and low-frequency coding variants alter human adult height”. In: Nature 2017 542:7640 542 (7640Feb. 2017), pp. 186–190. ISSN: 1476-4687.

[45] Adam E. Locke et al. “Exome sequencing of Finnish isolates enhances rare-variant association power”. In: Nature 2019 572:7769 572 (7769July 2019), pp. 323–328. ISSN: 1476-4687.

[46] Tomaz Berisa and Joseph K. Pickrell. “Approximately independent linkage disequilibrium blocks in human populations”. In: Bioinformatics 32 (2Jan. 2016), pp. 283–285. ISSN: 1367-4803.

