## Supplementary for "SUMMIT-FA: A new resource for improved transcriptome imputation using functional annotations"

| Phenotypes Analyzed by SUMMIT-FA | | |
|---|---|---|
| Asthma | Hypertension | Schizophrenia |
| Coronary Artery Disease | Hyperparathyroidism | Systemic Lupus Erythematosus |
| Diastolic Pressure | Irritable Bowel Disease | Systolic Pressure |
| Deep Vein Thrombosis | Insomnia | Type-I Diabetes |
| Eosinophil Count | Lymphocyte Count | Type-II Diabetes |
| Eczema | Monocyte Count | Triglycerides |
| Gout | Psoriasis | Ulcerative Colitis |
| High Cholesterol | Rheumatoid Arthritis | Waist Circumference |

Table 1: **24 phenotypes used in TWAS analysis.**

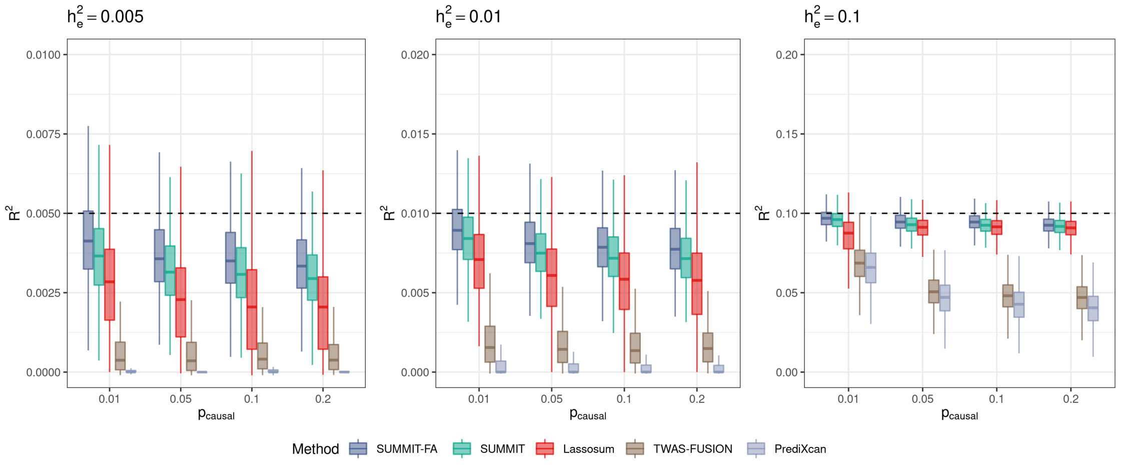


(a) $h_p^2 = 0.2$

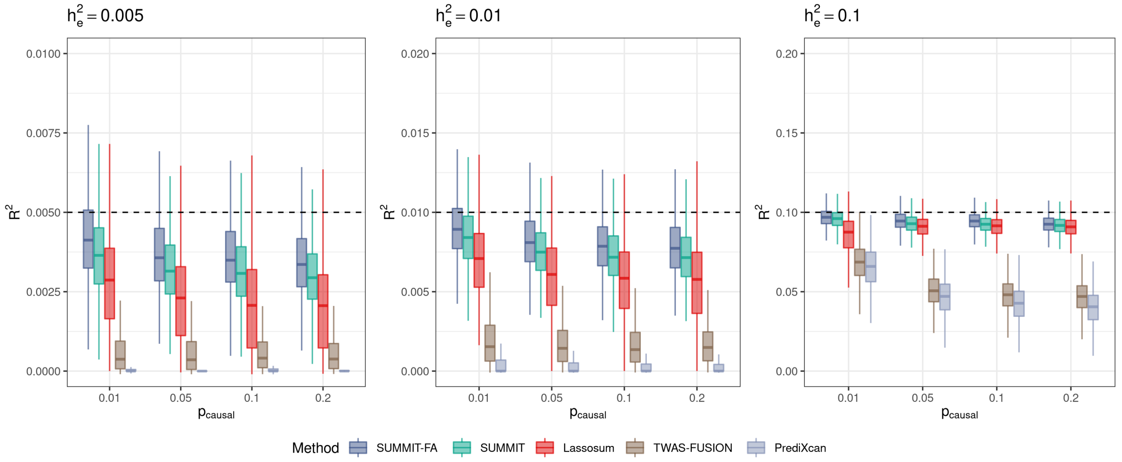


(b) $h_p^2 = 0.5$

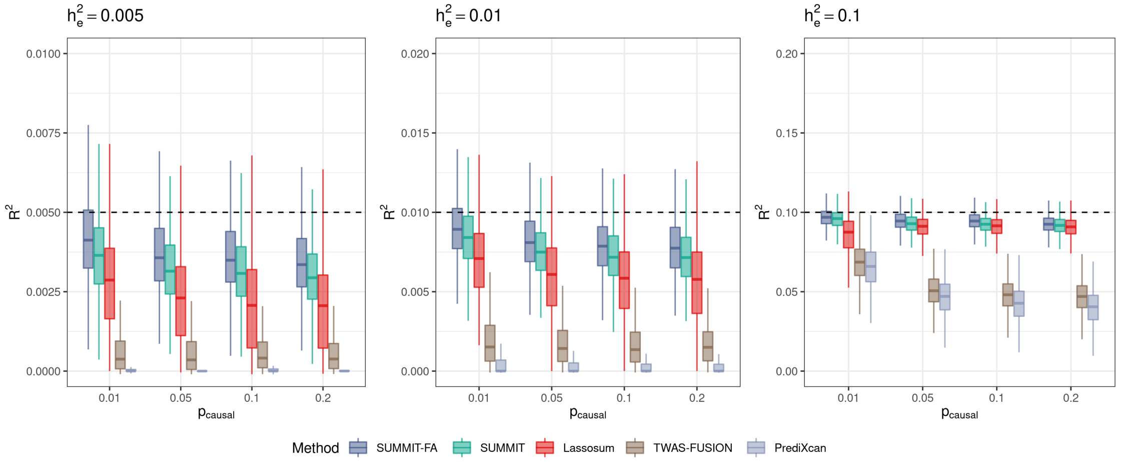


(c) $h_p^2 = 0.8$

Figure 1: **Simulation performance comparison based on gene *CHURC1*.** Plots of imputation $R^2$ in test samples by SUMMIT-FA, SUMMIT, Lassosum, TWAS-FUSION, and PrediXcan for varied proportion of causal SNPs $\mathrm{p}_{\mathrm{causal}}$, DNAm heritability $h_e^2$, and phenotypic heritability $h_p^2$.

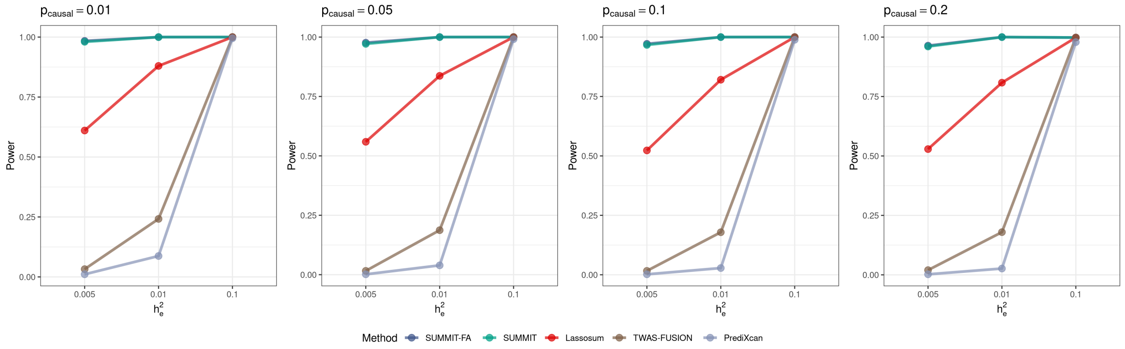


(a) $h_p^2 = 0.2$

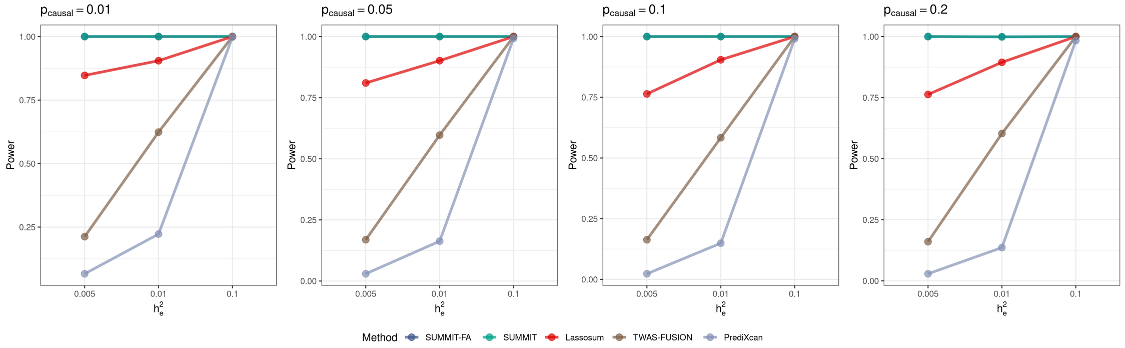


(b) $h_p^2 = 0.5$

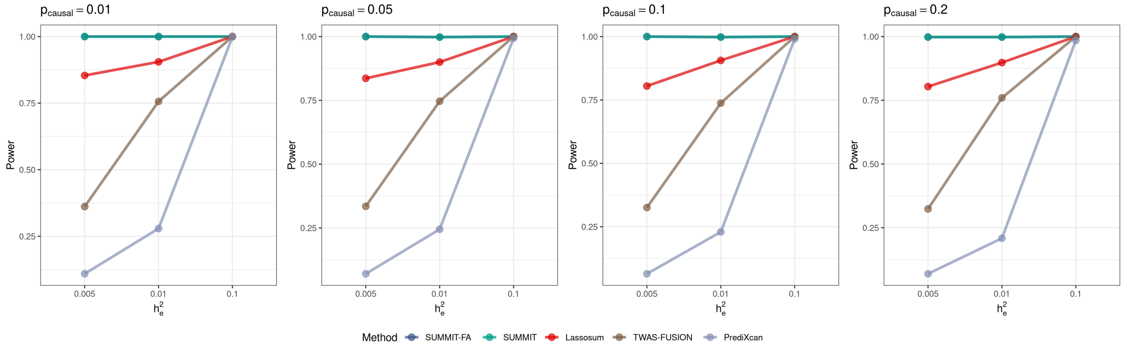


(c) $h_p^2 = 0.8$

Figure 2: **Simulation performance comparison based on gene *CHURC1*.** Plots of statistical power for SUMMIT-FA, SUMMIT, Lassosum, TWAS-FUSION, and PrediXcan for varied proportion of causal SNPs $\mathrm{p_{causal}}$, DNAm heritability $h_e^2$, and phenotypic heritability $h_p^2$. Empirical power was determined by the proportion of p-values $< 2.5 \times 10^{-6}$.

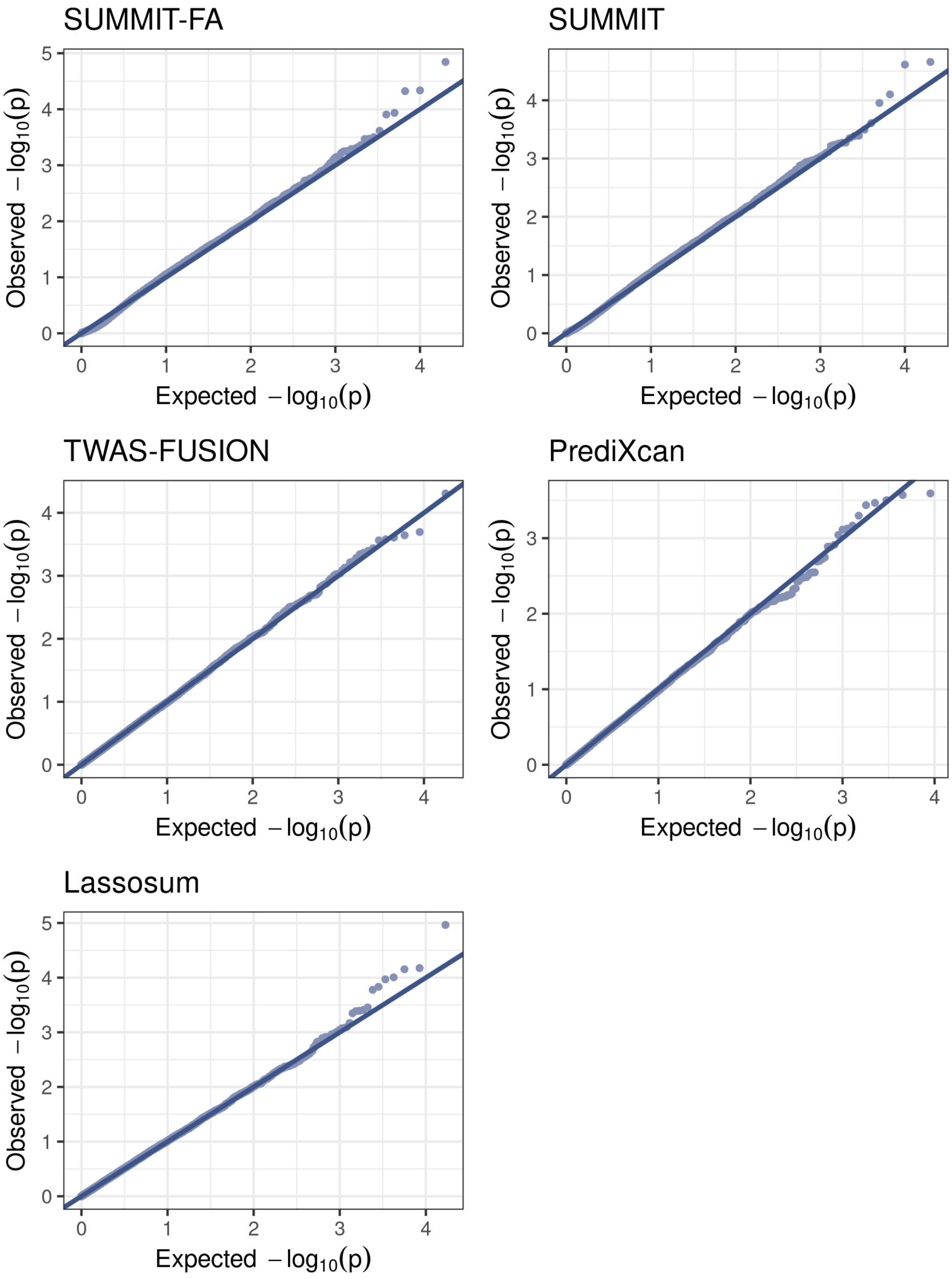


Figure 3: **QQ plots of p-values under $H_0$ based on the *CHURC1* gene.** 20,000 replicates were run under the null hypothesis to examine type I error rates, with $h_p^2 = 0.2$, $h_e^2 = 0.005$, $p_{\text{causal}} = 0.05$.

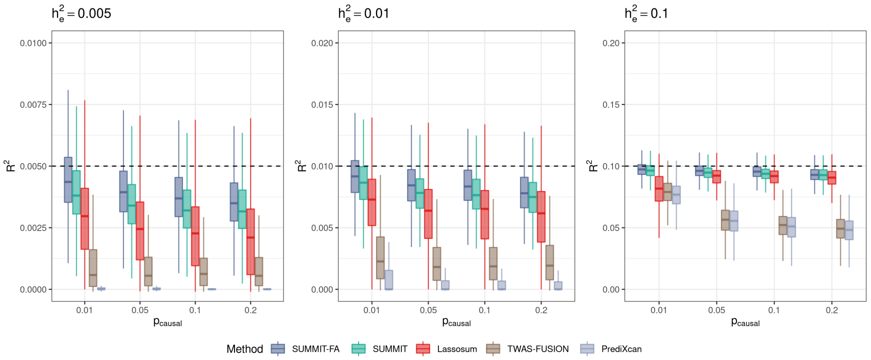


(a) $h_p^2 = 0.1$

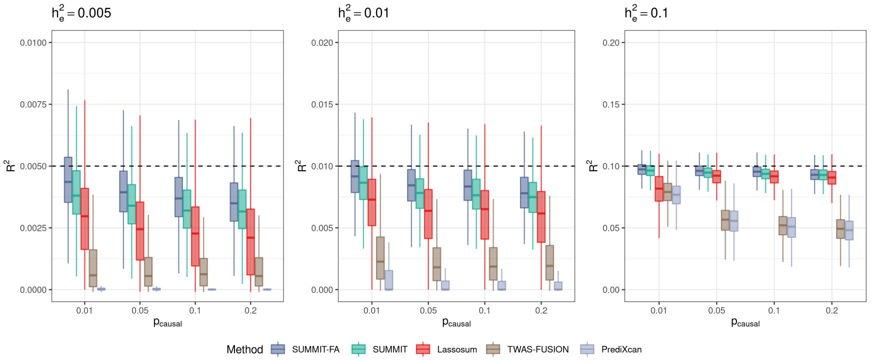


(b) $h_p^2 = 0.2$

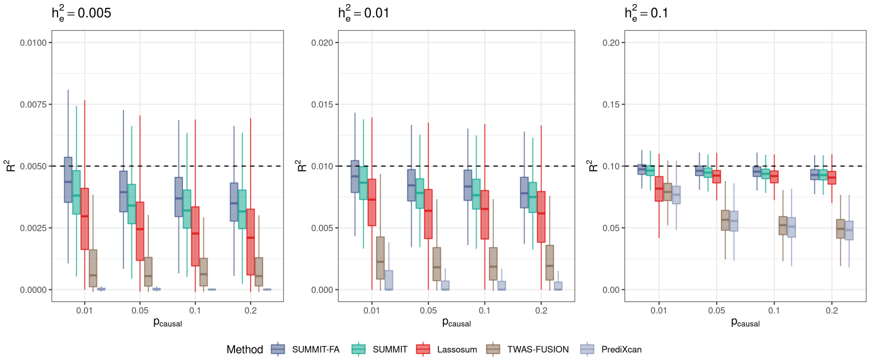


(c) $h_p^2 = 0.5$

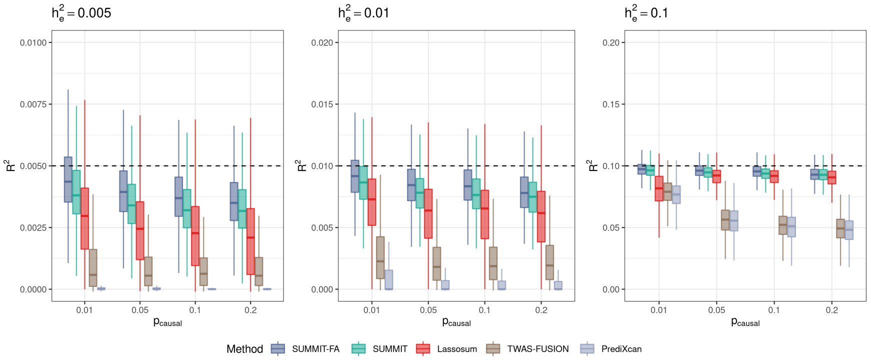


(d) $h_p^2 = 0.8$

Figure 4: **Simulation performance comparison based on gene *KRIT1*.** Plots of imputation $R^2$ in test samples by SUMMIT-FA, SUMMIT, Lassosum, TWAS-FUSION, and PrediXcan for varied proportion of causal SNPs $p_{causal}$, DNAm heritability $h_e^2$, and phenotypic heritability $h_p^2$.

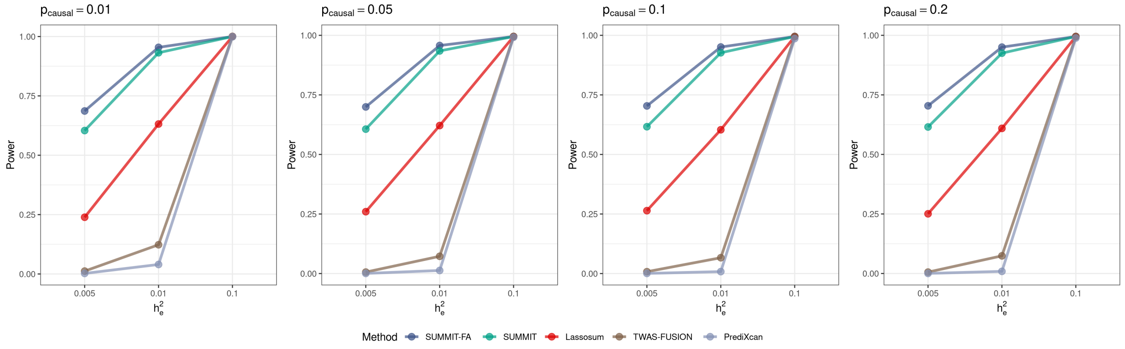


(a) $h_p^2 = 0.1$

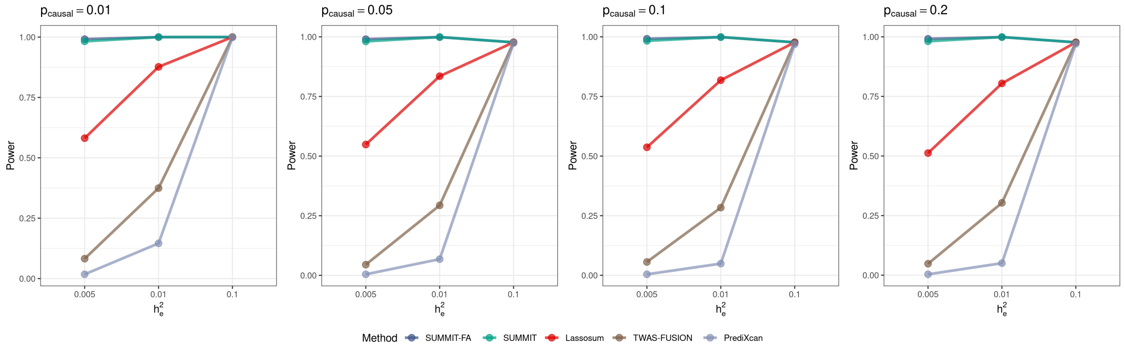


(b) $h_p^2 = 0.2$

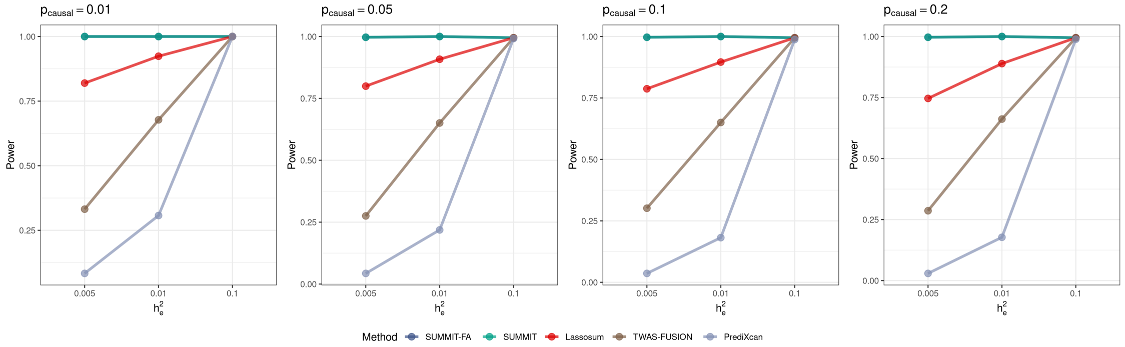


(c) $h_p^2 = 0.5$

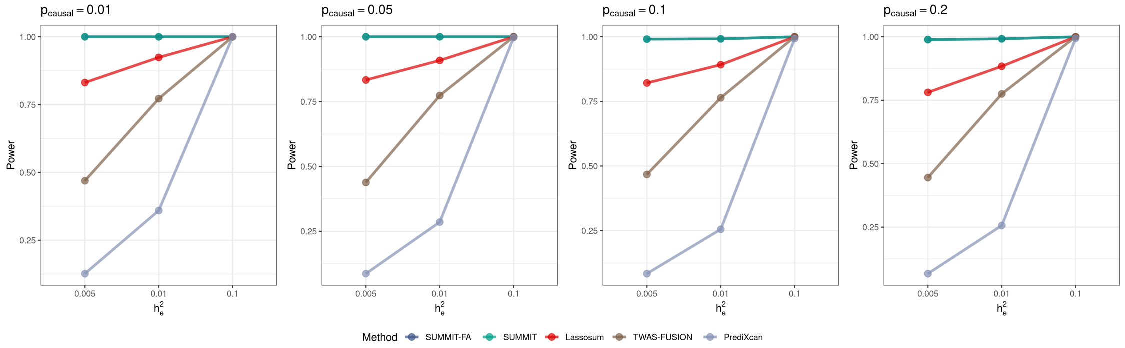


(d) $h_p^2 = 0.8$

Figure 5: **Simulation performance comparison based on gene *KRIT1*.** Plots of statistical power for SUMMIT-FA, SUMMIT, Lassosum, TWAS-FUSION, and PrediXcan for varied proportion of causal SNPs $p_{causal}$, DNAm heritability $h_e^2$, and phenotypic heritability $h_p^2$. Empirical power was determined by the proportion of p-values $< 2.5 \times 10^{-6}$.
